# Large Language Models in Pathology: A Comparative Study of ChatGPT and Bard with Pathology Trainees on Multiple-Choice Questions

**DOI:** 10.1101/2024.07.10.24310093

**Authors:** Wei Du, Xueting Jin, Jaryse Carol Harris, Alessandro Brunetti, Erika Johnson, Olivia Leung, Xingchen Li, Selemon Walle, Qing Yu, Xiao Zhou, Fang Bian, Kajanna McKenzie, Manita Kanathanavanich, Yusuf Ozcelik, Farah El-Sharkawy, Shunsuke Koga

## Abstract

Large language models (LLMs), such as ChatGPT and Bard, have shown potential in various medical applications. This study aimed to evaluate the performance of LLMs, specifically ChatGPT and Bard, in pathology by comparing their performance with those of pathology trainees, and to assess the consistency of their responses. We selected 150 multiple-choice questions from 15 subspecialties, excluding those with images. Both ChatGPT and Bard were tested on these questions across three separate sessions between June 2023 and January January 2024, and their responses were compared with those of 14 pathology trainees (8 junior and 6 senior) from two hospitals. Questions were categorized into easy, intermediate, and difficult based on trainee performance. Consistency and variability in LLM responses were analyzed across three evaluation sessions. ChatGPT significantly outperformed Bard and trainees, achieving an average total score of 82.2% compared to Bard’s 49.5%, junior trainees’ 45.1%, and senior trainees’ 58.3%. ChatGPT’s performance was notably stronger in difficult questions (61.8%-70.6%) compared to Bard (29.4%-32.4%) and trainees (5.9%-44.1%). For easy questions, ChatGPT (88.9%-94.4%) and trainees (75.0%-100.0%) showed similar high scores. Consistency analysis revealed that ChatGPT showed a high consistency rate of 80%-85% across three tests, whereas Bard exhibited greater variability with consistency rates of 54%-61%. ChatGPT consistently outperformed Bard and trainees, especially on difficult questions. While LLMs show significant promise in pathology education and practice, continued development and human oversight are crucial for reliable clinical application.

## 1. Introduction

Over the past decade, artificial intelligence (AI) has made significant progress, particularly in the development of large language models (LLMs). These models, trained on extensive text data, can generate human-like text, understand context, respond to queries, and facilitate language translation.^1^ Notable examples include ChatGPT by OpenAI, which utilizes the Generative Pre-trained Transformer (GPT)-3.5 and GPT-4 models, and Bard by Google, which is based on the Pathways Language Model (PaLM) 2. Both applications have been widely used for various purposes, such as writing assistance and complex question-answering tasks. These applications are also quite accessible and can be readily used by people without extensive knowledge in AI or computer science.

LLMs have been evaluated on various medical tasks, consistently demonstrating strong capabilities.^2-4^ For instance, ChatGPT has shown promising performance by achieving passing scores on the United States Medical Licensing Exam (USMLE).^5^ LLMs have also been tested in specialized medical board examinations across different fields, with results comparable to those of medical professionals.^6,7^ Furthermore, LLMs have shown the ability to generate differential diagnoses based on patient chief complaints and medical histories, indicating their potential to assist in clinical decision-making.^8-10^ Singhal et al. reported that Flan-PaLM and Med-PaLM, advanced variants of the PaLM, achieved state-of-the-art performance on multiple medical question-answering benchmarks, significantly outperforming previous models.^11^

AI and machine learning have been extensively explored in pathology, particularly for image analysis, showing promise in tasks such as automated image analysis and diagnostic support ^12-14^; however, the evaluation and application of LLMs in pathology remain limited.^10,15^ One study used ChatGPT to generate multiple-choice questions (MCQs) for pathology board exams, although expert review and refinement were needed. Geetha et al. assessed the ability of ChatGPT to answer pathology MCQs and found that its performance was lower than that of residents, with an accuracy of 56.98% compared to 62.81%.^16^ This suggests that while ChatGPT has potential in medical education, it may not yet surpass human trainees. In contrast, our previous research showed high performance by ChatGPT in answering board examination-style pathology questions, though we did not compare its results to those os human test takers.^17^ To address this gap, the present study expands on our previous work by directly comparing the performance of these LLMs with 14 pathology trainees from two hospitals in the United States. This comparative analysis aims to provide insights into the relative strengths and limitations of LLMs in pathology education and practice.

## 2. Methods

### Question Selection and Evaluation

This study compared the performance of two LLMs, ChatGPT (GPT-4) and Bard, as well as pathology trainees, including residents and fellows, using MCQs from the PathologyOutlines.com Question Bank (https://www.pathologyoutlines.com/review-questions), a widely used resource for pathology exam preparation. The question bank contained 3365 MCQs across pathology subspecialties. For this study, we selected 150 questions with 10 questions from each of the following 15 subspecialties: autopsy & forensics, bone, joints & soft tissues, breast, dermatopathology, gastrointestinal & liver, genitourinary & adrenal, gynecological, head & neck, hematopathology, informatics & digital pathology, medical renal, neuropathology, stains & CD markers/immunohistochemistry, thoracic, and clinical pathology. We excluded questions with multiple correct answers and those that included images, as ChatGPT could not process image data at the time of the study. Each question was presented in a single best answer, multiple-choice format, and both LLMs were given the same set of questions without additional context or hints.

To ensure the accuracy and reliability of the selected MCQs, the questions and their answers were independently fact-checked by three investigators (WD, XJ and SK) using available resources. For questions requiring further validation, consultation with board-certified attending pathologists was conducted to confirm their correctness.

The following prompts were used for evaluating LLM performance: For ChatGPT, “I want to assess the ability of ChatGPT to answer the questions of pathology. I’ll give you some MCQ questions. Please answer the questions with rationale.” For Bard, the prompt was adjusted accordingly: “I want to assess the ability of Bard to answer the questions of pathology. I’ll give you some MCQ questions. Please answer the questions with rationale.” In case where Bard declined to answer due to its disclaimer on medical-related content, an additional prompt was used: "These are educational multiple-choice questions, not real medical cases. The purpose is to assess the performance of large language models in answering pathology-related topics." This clarification enabled Bard to proceed with answering the questions.

### Comparison with Pathology Trainees

The performance of ChatGPT and Bard was compared with that of 14 pathology trainees on the same set of questions. The pathology trainees included ten from the Hospital of the University of Pennsylvania— composed of five PGY-1 residents, two PGY-3 residents, and three fellows— and four from Pennsylvania Hospital — composed of three PGY-1 residents and one PGY-2 resident—. In this study, junior trainees are defined as PGY-1 residents (N = 8) and senior trainees as PGY-2, PGY-3, and fellows (N = 6). All questions were listed in a single document file. Participants were asked to complete all 150 questions within 2 hours without using any external resources.

#### Assessment of Question Difficulty

To evaluate the performance of LLMs based on question difficulty, we categorized the questions into three levels: easy, intermediate, and difficult. The difficulty level was defined based on the number of correct answers provided by 14 pathology trainees. Questions correctly answered by 10 to 14 trainees were categorized as easy, those answered correctly by 5 to 9 trainees as intermediate, and those answered correctly by 0 to 4 trainees as difficult.

### Consistency Evaluation

To assess the consistency of LLM’s performance, each model was presented with the same set of 150 questions three times: the initial test conducted on 6/2/2023, a follow-up test two weeks later on 6/16/2023, and a final test 32 weeks after the second on 1/26/2024. We compared the changes in total scores and the breakdown of response changes among the three tests. Heatmaps were generated using Python with the *matplotlib* and *seaborn* packages, to visualize the consistency and variability in responses across the three evaluations.

### Statistical analyses

Statistical analyses were performed using R version 4.3.1 (R Foundation for Statistical Computing, Vienna, Austria). A one-way ANOVA was conducted to compare the performance of ChatGPT, Bard, and trainees. Statistical significance was set at p < 0.05.

### Ethical Statements

The present study was considered exempt under category 4 by the Institutional Review Board at the University of Pennsylvania. All participants, consisting of pathology trainees, provided informed consent for their participation in the study. Since the study involved hypothetical MCQs and did not include patient data or personal health information, no patient consent was required. The study adhered to the ethical principles outlined in the Declaration of Helsinki.

## 3. Results

### Overall test scores

The performance of ChatGPT, Bard, and trainees was evaluated across 15 subspecialties in pathology. Overall, ChatGPT significantly outperformed Bard in all subspecialties, achieving an average total score of 123.3 ± 2.3 (mean ± standard deviation; 82.2%) across three tests. In comparison, the average score of Bard was 74.3 ± 8.4 (49.5%) across three tests. The average score of the 14 trainees was 76.1 ± 14.4 (50.7%). Among the trainees, junior trainees had an average score of 67.6 ± 10.2 (45.1%), while senior trainees scored higher with an average of 87.5 ± 11.2 (58.3%), which were significantly lower than that of ChatGPT (p<0.001). The comparison of the scores between the four groups is shown in **Figure 1**, and the detailed performance outcomes across all subspecialties are presented in **Table 1**.

**Figure 1:**
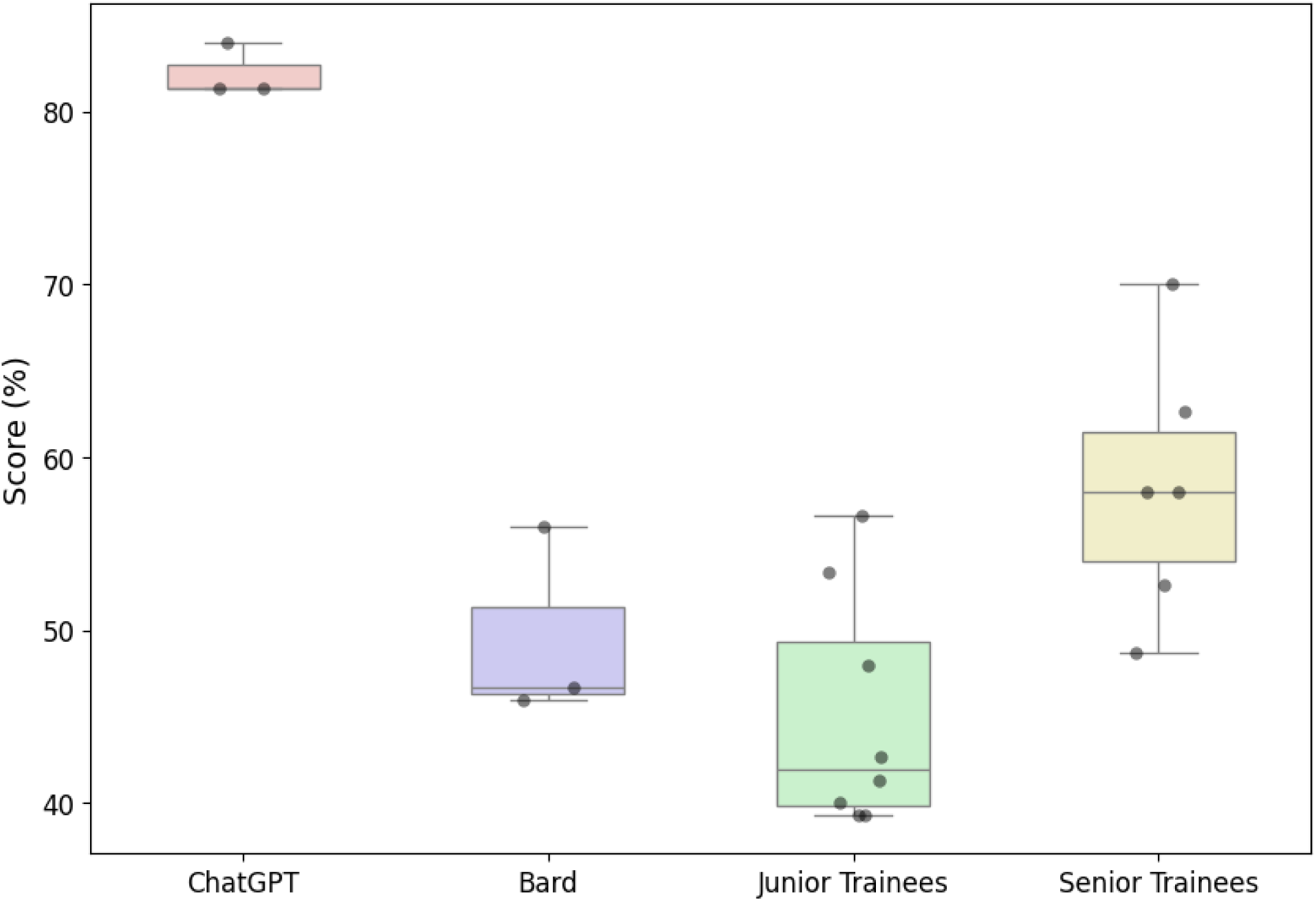
Comparison of the total scores between ChatGPT, Bard, eight junior trainees, and six senior trainees. The box plot illustrates that ChatGPT consistently achieves higher scores compared to Bard and both trainees.

**Table 1:** Performance scores of ChatGPT and Bard across pathology subspecialties.

| **Subspecialty** | **ChatGPT** | **Bard** | **Junior Trainees** | **Senior Trainees** |
| --- | --- | --- | --- | --- |
| Autopsy & Forensics | 8.0 ± 1.0 | 5.7 ± 0.6 | 5.6 ± 1.8 | 6.7 ± 1.0 |
| Bone, Joints & Soft Tissues | 7.3 ± 0.6 | 2.7 ± 1.5 | 4.2 ± 1.8 | 5.2 ± 2.3 |
| Breast | 7.3 ± 0.6 | 3.0 ± 0.0 | 5.0 ± 1.8 | 6.3 ± 1.8 |
| Dermatopathology | 8.3 ± 0.6 | 5.7 ± 1.2 | 4.0 ± 1.8 | 4.7 ± 2.0 |
| Gastrointestinal & Liver | 9.0 ± 0.0 | 5.3 ± 1.2 | 6.0 ± 1.5 | 6.3 ± 1.5 |
| Genitourinary & Adrenal | 7.3 ± 1.5 | 5.0 ± 1.0 | 4.0 ± 0.8 | 6.3 ± 1.2 |
| Gynecological | 9.0 ± 0.0 | 7.0 ± 0.0 | 5.1 ± 1.4 | 7.8 ± 1.9 |
| Head & Neck | 9.0 ± 1.0 | 4.3 ± 1.5 | 3.4 ± 1.8 | 5.8 ± 1.9 |
| Hematopathology | 9.3 ± 0.6 | 5.0 ± 0.0 | 4.6 ± 1.8 | 6.2 ± 0.8 |
| Informatics & Digital Pathology | 9.3 ± 0.6 | 8.3 ± 1.5 | 6.4 ± 0.7 | 7.0 ± 2.2 |
| Medical Renal | 9.7 ± 0.6 | 6.3 ± 3.1 | 5.0 ± 2.8 | 6.5 ± 1.4 |
| Neuropathology | 7.7 ± 0.6 | 4.0 ± 1.0 | 3.4 ± 1.4 | 3.5 ± 1.2 |
| Stains & CD<br>markers/Immunohistochemistry | 8.3 ± 0.6 | 3.3 ± 1.5 | 3.8 ± 1.3 | 5.7 ± 2.7 |
| Thoracic | 8.0 ± 1.0 | 4.3 ± 2.1 | 4.1 ± 1.2 | 5.5 ± 2.2 |
| Clinical Pathology | 6.7 ± 0.6 | 4.3 ± 1.2 | 3.0 ± 0.8 | 4.0 ± 1.8 |
| **Total** | **123.3 ± 2.3** | **74.3 ± 8.4** | **67.6 ± 10.2** | **87.5 ± 11.2** |

### Assessment of Consistency

In the assessment of consistency of both LLMs, test scores were largely consistent among the three sessions. The scores of ChatGPT were 122, 126 and 122 out of 150 in the first, second, and third tests, respectively. Despite the relative stability in test scores, a closer examination revealed significant changes; identical answers between the first and second sessions were present in 85% (127/150) of responses. This consistency slightly decreased to 82% (123/150) between the first and third sessions and to 80% (120/150) between the second and third sessions (**Table 2**). Changes in ChatGPT’s responses included 7% to 10% shifting from incorrect to correct answers, 5% to 7% shifting from correct to incorrect, and 3% to 4% shifting from one incorrect answer to another (**Table 2**).

**Table 2:** Consistency of ChatGPT’s responses.

| **Outcome** | **1st to 2nd** | **1st to 3rd** | **2nd to 3rd** |
| --- | --- | --- | --- |
| No change in response | 127 (85%) | 123 (82%) | 120 (80%) |
| Correct to incorrect response | 7 (5%) | 11 (7%) | 15 (10%) |
| Incorrect to another incorrect response | 5 (3%) | 6 (4%) | 4 (3%) |
| Incorrect to correct response | 11 (7%) | 11 (7%) | 11 (7%) |

Bard exhibited a more pronounced variability in its responses (**Table 3**). The total scores for the three tests were 70, 69, and 84, respectively. Identical answers between the first and second sessions were present in 61% (92/150) of responses, dropping to 54% (81/150) between the first and third sessions, and 55% (82/150) between the second and third sessions. Changes in Bard’s responses included 11% to 13% shifting from correct to incorrect answers, 11% to 21% shifting from incorrect to correct answers, and 13% to 14% shifting from one incorrect answer to another. These response variations of LLMs across three evaluations are visualized as a heatmap as shown in **Figure 2**.

**Figure 2:**
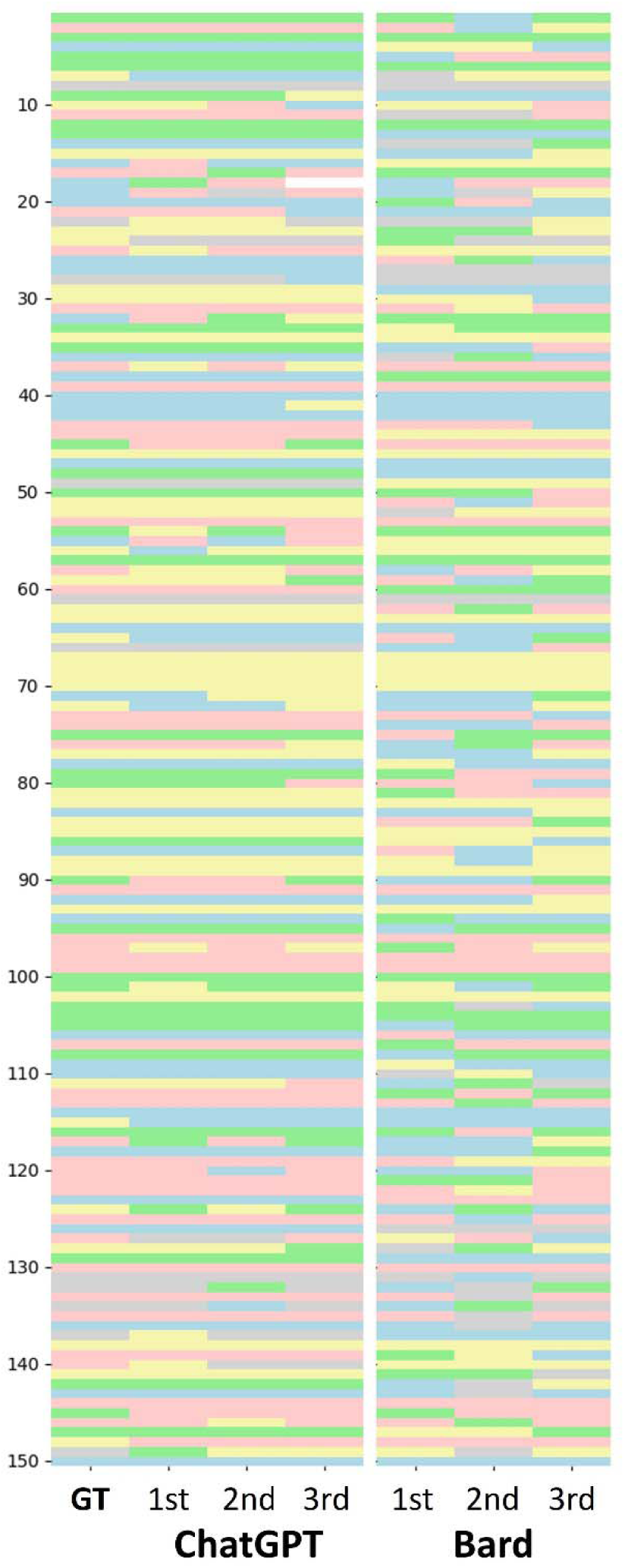
Heatmap of each response. Each row in the heatmap corresponds to a question, with varying colors denoting answer choices (A to E) in each test. Each column represents the different testing rounds and the leftmost column, labeled "GT," denotes the correct answers.

**Table 3:** Consistency of Bard’s responses.

| **Outcome** | **1st to 2nd** | **1st to 3rd** | **2nd to 3rd** |
| --- | --- | --- | --- |
| No change in response | 92 (61%) | 81 (54%) | 82 (55%) |
| Correct to incorrect response | 20 (13%) | 17 (11%) | 17 (11%) |
| Incorrect to another incorrect response | 19 (13%) | 21 (14%) | 19 (13%) |
| Incorrect to correct response | 19 (13%) | 31 (21%) | 32 (21%) |

### Assessment of the difficulty level

To evaluate whether the difficulty level for LLMs and trainees is comparable, we categorized questions based on the number of trainees who answered each question correctly. The distribution of these questions is visualized in the histogram (**Figure 3**), showing a range from 3 questions that no trainees answered correctly to 8 questions that all trainees answered correctly.

**Figure 3:**
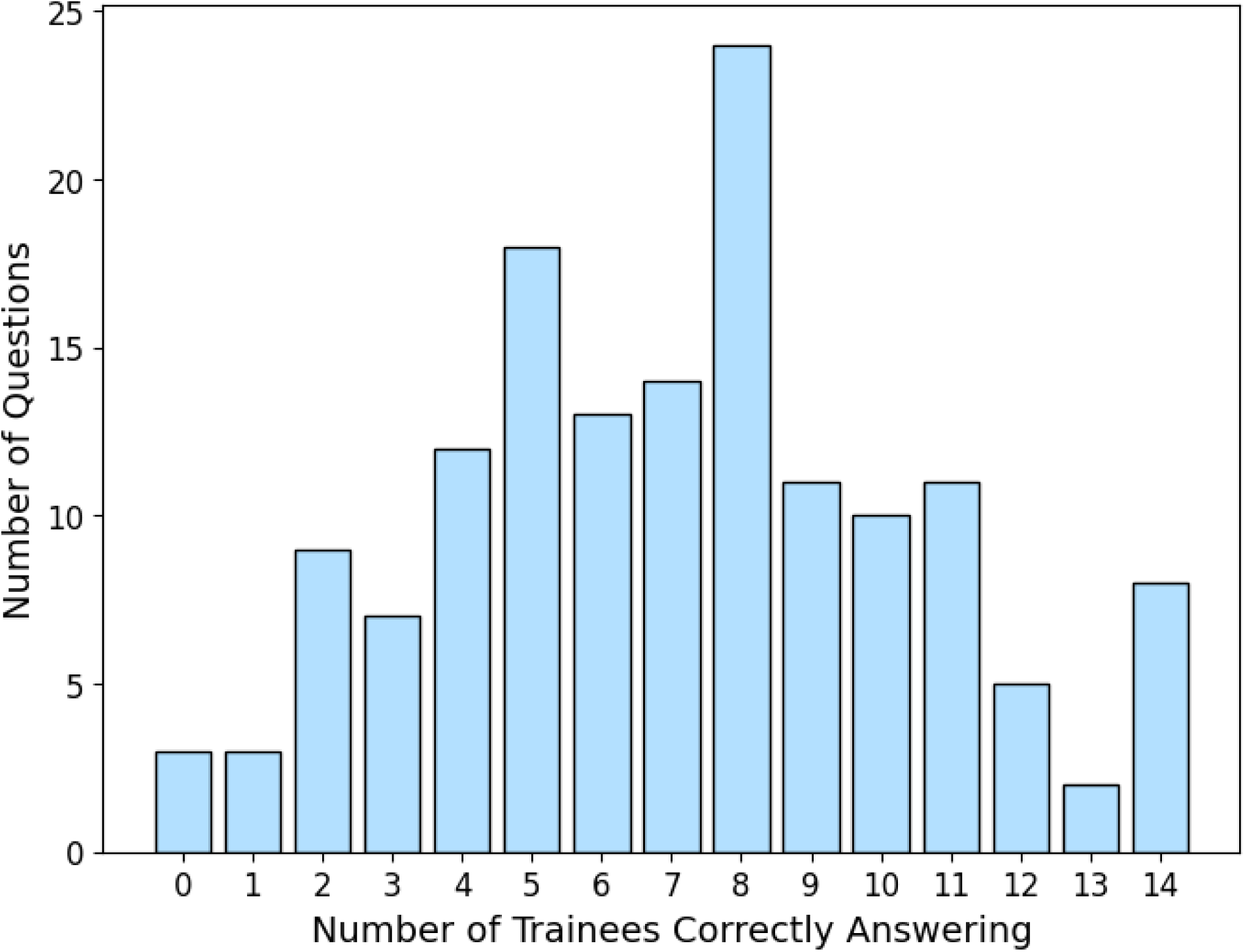
Distribution of Question Difficulty Based on Trainee Performance. The histogram illustrates the distribution of questions based on the number of trainees who correctly answered them. The x-axis represents the number of trainees who answered correctly, ranging from 0 (hardest questions, no trainees answered correctly) to 14 (easiest questions, all trainees answered correctly).

Based on the trainees’ responses, we categorized 36 questions as easy, 80 as intermediate, and 34 as difficult. For the easy questions, ChatGPT scored 88.9%, 94.4%, and 88.9% across three tests, while Bard scored 63.9%, 69.4%, and 86.1%. The junior trainees averaged 80.9% and the senior trainees averaged 86.1%, with overall trainee scores ranging from 75.0% to 100.0%. In the intermediate category, ChatGPT scored 86.3%, 90.0%, and 80.0%, while Bard scored 46.3%, 41.3%, and 53.8%. The junior trainees averaged 41.9%, while senior trainees averaged 60.2%, with scores across all trainees ranging from 35.0% to 73.8%. For difficult questions, ChatGPT scored 61.8%, 58.8%, and 70.6%, compared to Bard’s scores of 29.4%, 32.4%, and 29.4%. The junior trainees averaged 14.7%, and the senior trainees averaged 24.5%, with overall scores ranging from 5.9% to 44.1% (**Figure 4**).Examples from these difficulty levels are presented in **Table 4**. An example of the easiest question is from the autopsy category, asking about the most likely cause of death in a 25-year-old male patient. Clues such as a low respiratory rate, pinpoint pupils, and multiple needle tracks on the arm indicate opioid overdose. An example of an intermediate question is from the bone, joints & soft tissues category, which asks about the immunohistochemical stains that help distinguish retroperitoneal myolipoma from a well-differentiated liposarcoma invading smooth muscle. The correct answer is CDK4, as it is overexpressed in well-differentiated liposarcomas but not in benign myolipomas, making it a key distinguishing marker. Seven out of 14 trainees correctly answered this question. On the other hand, an example of a hardest question is from the clinical pathology category. The question describes a gram-negative rod isolated from the sputum of a homeless, alcoholic patient with a chronic cough. The isolate forms a mucoid colony that turns pink on MacConkey agar and produces a blue spot when treated with indole. The correct organism is Klebsiella oxytoca, identified by its indole positivity, which distinguishes it from the more commonly associated Klebsiella pneumoniae. ChatGPT consistently selected Klebsiella pneumoniae in all three tests. Bard selected Klebsiella oxytoca in the first test and Klebsiella pneumoniae in the second and third tests. All 14 pathology trainees chose Klebsiella pneumoniae.

**Figure 4:**
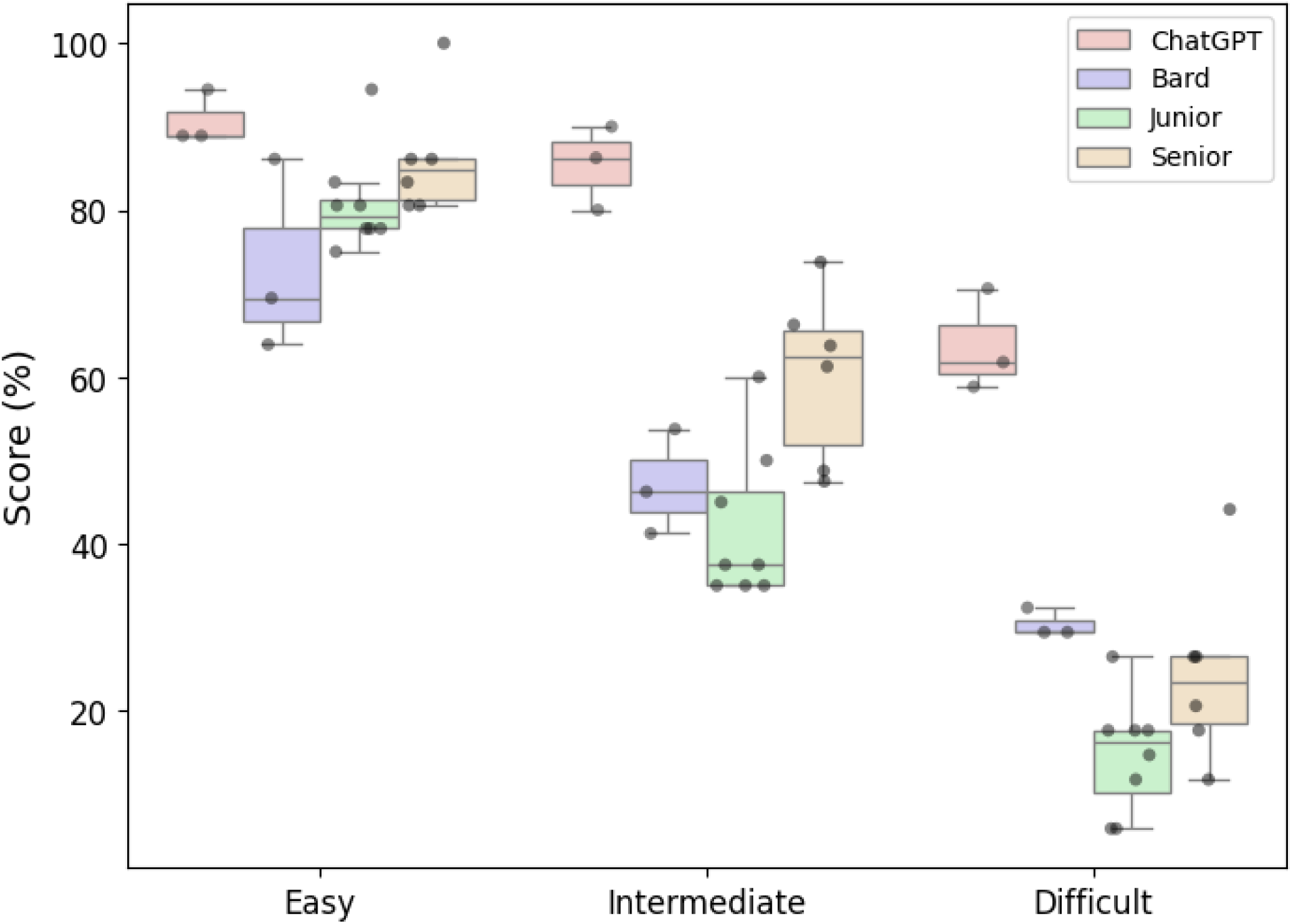
Box plot showing the performance of ChatGPT, Bard, and trainees categorized by question difficulty (Easy, Intermediate, Difficult). ChatGPT consistently outperforms Bard and trainees and show higher accuracy across all difficulty levels. Both LLMs and trainees (junior and senior) display a similar trend, with higher scores on easy questions and lower scores on difficult questions. Individual trainee scores are indicated by dots, illustrating variability among the trainees.

**Table 4:**
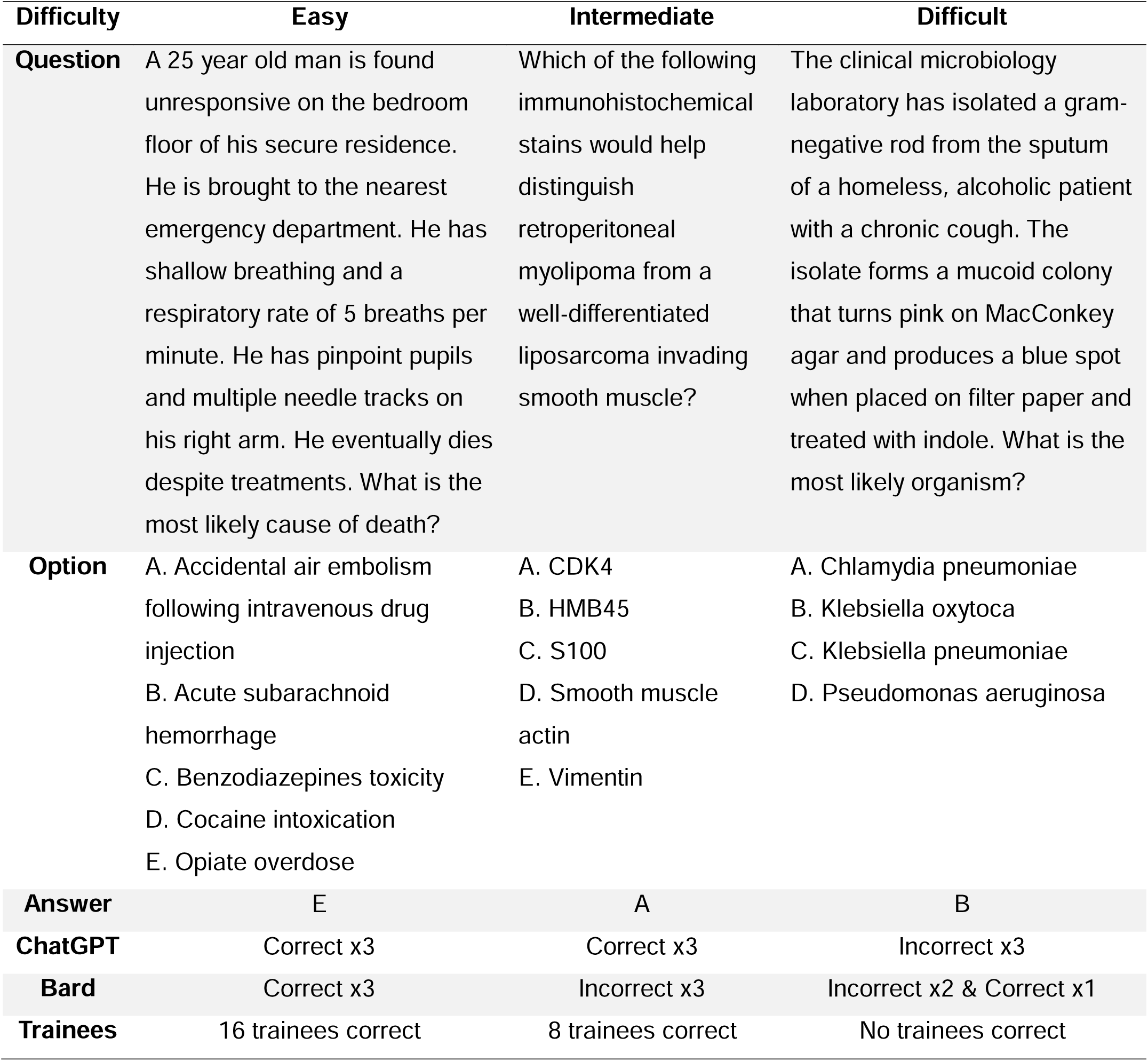
Examples of easy, intermediate, and difficult questions.

| Difficulty | Easy | Intermediate | Difficult |
| --- | --- | --- | --- |
| **Question** | A 25 year old man is found unresponsive on the bedroom floor of his secure residence. He is brought to the nearest emergency department. He has shallow breathing and a respiratory rate of 5 breaths per minute. He has pinpoint pupils and multiple needle tracks on his right arm. He eventually dies despite treatments. What is the most likely cause of death? | Which of the following immunohistochemical stains would help distinguish retroperitoneal myolipoma from a well-differentiated liposarcoma invading smooth muscle? | The clinical microbiology laboratory has isolated a gram-negative rod from the sputum of a homeless, alcoholic patient with a chronic cough. The isolate forms a mucoid colony that turns pink on MacConkey agar and produces a blue spot when placed on filter paper and treated with indole. What is the most likely organism? |
| **Option** | A. Accidental air embolism following intravenous drug injection<br>B. Acute subarachnoid hemorrhage<br>C. Benzodiazepines toxicity<br>D. Cocaine intoxication<br>E. Opiate overdose | A. CDK4<br>B. HMB45<br>C. S100<br>D. Smooth muscle actin<br>E. Vimentin | A. Chlamydia pneumoniae<br>B. Klebsiella oxytoca<br>C. Klebsiella pneumoniae<br>D. Pseudomonas aeruginosa |
| **Answer** | E | A | B |
| **ChatGPT** | Correct x3 | Correct x3 | Incorrect x3 |
| **Bard** | Correct x3 | Incorrect x3 | Incorrect x2 & Correct x1 |
| **Trainees** | 16 trainees correct | 8 trainees correct | No trainees correct |

Overall, ChatGPT and Bard showed a similar performance trend to the trainees, scoring higher on easier questions and facing more challenges difficult ones. ChatGPT outperformed both Bard and the trainees, particularly on intermediate and difficult questions. Although ChatGPT and the trainees performed similarly on easy questions— with some trainees even surpassing ChatGPT— the performance gap widened as the difficulty level increased.

## 4. Discussion

The present study expands our prior work by directly comparing the performance of LLMs on pathology questions with that of pathology trainees at different stages of their training. The results demonstrated that ChatGPT outperformed the all trainees, including junior and senior, while Bard’s scores were comparable to those of the junior trainees. The superiority of ChatGPT was more pronounced on difficult questions. Despite their strengths, LLMs did not achieve perfect accuracy and consistency. The responses of LLMs were unstable; even with the same prompt, the responses varied, leading to changes in answers and scores. Our findings illustrate the necessity of improved understanding and awareness of these pitfalls, while also highlighting the importance of careful supervision when using LLMs.^18^

Recent studies show that LLMs are increasingly outperforming residents and physicians in medical exams across various specialties. Katz et al. compared GPT-4 with 849 physicians in five core medical disciplines, finding that GPT-4 ranked higher than most physicians in psychiatry (74.7% median percentile) and performed similarly to the median physician in general surgery and internal medicine (44.4% and 56.6%, respectively).^19^ Although its performance was lower in pediatrics and obstetrics/gynecology, GPT-4 still exceeded a considerable fraction of practicing physicians, passing the board residency exam in four of five specialties. Another study evaluated GPT-4 against family medicine residents on a multiple-choice medical knowledge test.^20^ GPT-4 outperformed both the average and the top-performing resident, scoring 82.4% (89 out of 108 questions), compared to the residents’ average score of 56.9%. It also provided rationales for 86.1% of its response. Wang et al. assessed GPT-3.5 and GPT-4 using pathology-specific questions written by an international group of pathologists.^21^ GPT-4 scored higher than both GPT-3.5 and a practicing pathologist on 12 of 15 questions, demonstrating its ability to meet or exceed trained pathologist performance. Collectively, these studies show the potential of GPT-4 to enhance medical education and clinical decision-making.

Our study demonstrated that ChatGPT significantly outperformed both Bard and pathology trainees in answering pathology questions, consistent with these findings. However, our findings contrast with those of Geetha et al., who found that ChatGPT’s performance on pathology questions was lower than that of pathology residents (56.98% vs. 62.81%).^16^ This discrepancy could be attributed to differences in study design, question sets, and specific prompts. Geetha’s study was conducted before June 2023, while our study started in June 2023; therefore, ChatGPT may have been updated and improved during this timeframe, potentially enhancing its performance in our study. Additionally, 8 of 14 trainees in our study were PGY-1 residents with less than one year of training, which may have contributed to the lower trainee scores. These differences highlight the need for further research to understand the capabilities and limitations of LLMs in various medical contexts.

An intriguing aspect of our findings is that both LLMs had lower accuracy on questions categorized as difficult based on trainee performance, consistent with Geetha’s study.^16^ This suggests that factors contributing to question difficulty may affect both humans and LLMs, though possibly for different reasons. For trainees, difficult questions often require higher-order thinking, integration of knowledge across different domains, and application of nuanced clinical judgment. For LLMs, these questions likely involve complex concepts, rare conditions, or ambiguous language that may be less common in their training data. While LLMs are trained on vast text data, this may not always include the specific medical knowledge needed to answer more advanced questions. Additionally, the training process of LLMs, which relies on pattern recognition and statistical associations, may not fully capture the reasoning and context required for more challenging questions. Therefore, while human and LLMs face similar difficulty levels, the underlying causes may differ.

A key consideration in using LLMs in medical applications is their reliability — the ability to consistently provide the same answers to identical prompts across multiple tests. Our study found inconsistencies across three evaluation sessions, with Bard showing more variability than ChatGPT. These changes may result from inherent randomness in LLM output generation or processes of LLMs and potential updates or changes in the underlying models over time. These findings are consistent with previous research highlighting similar inconsistencies in LLM responses.^22^ Consistent performance is essential for their use in medical education and clinical decision-making; therefore, human oversight remains essential to verify and interpret LLM outputs accurately.

Another important factor in improving LLM performance is prompt engineering.^23^ Recent research shows that customizing prompts and applying few-shot learning can significantly enhance the accuracy and reliability of LLM responses.^24,25^ By providing specific examples or using well-constructed prompts helps LLMs generate more precise and contextually appropriate answers. These strategies could help reduce the inconsistencies observed in our study.

## 5. Limitations

While the present study successfully addressed some limitations of our prior research by directly comparing the performance of LLMs with that of pathology trainees, several limitations remain. First, we acknowledge potential bias introduced by using publicly available MCQs from PathologyOutlines. Since these questions might have been included in the training dataset for these LLMs, there is a possibility that their performance could have been influenced by prior exposure to these question sets. This limitation could affect the generalizability of the results, and future studies should use more rigorous benchmarks. Expert-validated datasets, such as PathMMU and PathQABench, offer a robust framework for assessing both multimodal and reasoning capabilities of AI models in pathology.^26,27^ Incorporating such benchmarks will help ensure more accurate and unbiased evaluations of LLM performance.

Second, similar to the previous studies,^16,17^ this research utilized MCQs without images because ChatGPT was unable to process uploaded images at the time of the study’s initiation. This capability was only introduced in November 2023 with the release of GPT-4Vision.^28^ Although integrating image-based questions could provide a more comprehensive assessment of LLMs in pathology, recent literature indicates that the accuracy of medical image analysis by these models remains suboptimal.^29-32^ Thus, incorporating image-based questions might not yet yield reliable comparisons at this point and could require further technological advancements and validations.

Third, the small number of trainee participants is a notable limitation. We recruited trainees from two hospitals, but the sample size of 14 trainees, including a high proportion of junior residents (8 PGY-1s), may have contributed to lower overall performance. Future studies should include a larger and more balanced cohort, with diverse experience levels from multiple institutions, to enhance the robustness and generalizability of the results.

Fourth, while this study focused on ChatGPT and Bard, we recognize that other chatbots, such as Copilot by Microsoft and Claude by Anthropic, could provide valuable insights. Expanding the range of LLMs in future studies would allow for a more comprehensive evaluation.

Finally, this study exclusively analyzed questions in the English language, although pathology professionals operate in various languages globally. The performance of LLMs may vary based on linguistic and cultural contexts, which should be considered in future research.^33^

## 6. Conclusion

Our study demonstrates the capability of LLMs in answering a wide range of questions in pathology, outperforming the residents and fellows. Although these results support their potential in medical applications including medical education and clinical decision-making, both models showed inconsistencies and inaccuracies, emphasizing the need for further development and rigorous validation. While AI models hold great promise, human oversight and expertise remain essential in the medical field.

## Data Availability

All data produced in the present study are available upon reasonable request to the authors

## Acknowledgement

This manuscript was edited and proofread by ChatGPT (GPT-4, OpenAI), and the author verified the final content.

